# Clinical Pattern of Retinal Detachment at the University of Gondar Tertiary Eye Care and Training center, North-West Ethiopia

**DOI:** 10.1101/2022.06.21.22276702

**Authors:** Hagos Kidanemariam, Asamere Tsegw

## Abstract

**Objective:** Due to increment in volume of cataract surgery and diabetic retinopathy, the incidence of retinal detachment (RD) is expected to rise in the future. However, little is known about the clinical patterns and risk factor of RD in Ethiopia. This study was aimed to determine the clinical pattern and risk factors of RD at University of Gondar Tertiary eye care and training center (UOG-TECTC).

**Methods:** A hospital based prospective cross sectional study was done involving all consecutive patients with retinal detachment seen over six months period at the UOG-TECTC. Structured questionnaire was used to extract the required data which included demography, ophthalmic history and clinical examination results. Data were entered into SPSS version 20 and analyzed.

**Results:** A total of 73 eyes of 65 patients with RD were analyzed. Two-third of patients were males 42(64.6%) and the median age was 49 years. Rhegmatogenous RD (RRD) was the commonest type of RD found in 58(89.2%) patients. Three quarter of the patients, 49 (75 %), came after 1 month of the symptom occurrence; out of which 20 (41%) visited the center after 1year duration. The commonest identified risk factors were trauma 19(29%) followed by pseudophakia 11(17%). More than three-quarter of the patients 53(81.5%) had detached macula and 60 (92.3%) of the involved eyes were blind at presentation.

**Conclusion and recommendations:** RRD was the commonest type of RD seen in our patients. Majority of the patients visited our center late after macular detachment and trauma was the commonest risk factor. People should be educated to seek health care immediately after the onset of symptoms. Ocular safety precautions should be advised to decrease the risk of ocular trauma. The center should be equipped to manage patients with RD.

## Introduction

Retinal detachment (RD) is separation of the neurosensory retina from the retinal pigment epithelium. There are three clinical types of retinal detachments. These are rhegmatogenous, tractional and exudative of which rhegmatogenous detachment being the most common type. (1,2)

A retrospective study in Nigeria showed that RD accounts for 4.5% of all vitreoretinal disease and a prospective hospital based study in Jimma, Ethiopia showed that the hospital prevalence of blindness from RD was 1.5 %. (3, 2, 4)

In the developing countries, most patients come late with detachment of the macula in 82-90% of the cases. Retinal detachment is treatable if detected early but patients get blind because of delayed presentation. Reports from Nigeria and Ethiopia showed that 79.8% and 64.9% patients’ eyes with retinal detachment were blind at presentation respectively. (1, 2, 5)

The risk factors for retinal detachment may differ based on age and type of detachment; but cataract surgery, trauma, myopia, posterior uveitis, proliferative diabetic retinopathy,, lattice degeneration, end stage renal disease and bacterial endophthalmitis are some of the commonly reported risk factors. (6, 7, 8)

As the cataract surgical volume and the prevalence of Diabetic retinopathy increases in the developing countries, the prevalence RD is also expected to increase. So this study is designed to fill the gap in terms of determining the risk factors, severity and patterns of retinal detachment at the University of Gondar tertiary eye care and training center, Gondar, Ethiopia

## Patients and Methods

### Study Design and Period

A Hospital based prospective cross sectional study was conducted at the University of Gondar tertiary eye care and training center involving all new patients with retinal detachment from September 2018 to February 2019.

### Study Area

The study was conducted at University of Gondar Tertiary Eye Care and Training Center, a major eye care and training center in Ethiopia. It is an ophthalmic referral center for an estimated 14 million people living in North-West Ethiopia. The center provides eye care services both at base hospital and rural outreach sites and annually, over 80,000 patients are seen at both sites. The base hospital has 8 out-patient clinics, facilities for in-patient care with 30 beds and five operation theatres.

Available Facilities and equipment for the management of vitreoretinal diseases at the center include, vitreoretina out-patient clinic, a vitreoretinal consultant, Retina Laser treatment room with Diod laser and Argon Green Laser equipments, a Fundus Camera machine, four indirect ophthalmoscopes, B-scan ultrasound, OCT and Facilities for intravitreal pharmacotherapy (Steroid and Anti-VEGF). There are no equipment for Scleral buckling and posterior vitrectomy surgeries to manage RD.

### Study Population

All new patients with retinal detachment who visited the center during the study period were included in the study. Follow up cases were excluded from the study.

### Data collection tools and procedures

### DATA COLLECTION PROCEDURE AND QUALITY CONTROL

Structured interviewer-administered questionnaire, document review and ocular examination were used to collect data. The questionnaire consisted of three sections: Socio-demographic variables, ophthalmic history and checklist for clinical data extraction (five items). Data quality was ensured through pre-testing the questionnaire before the actual data collection period.

Comprehensive Ophthalmic evaluation which included detailed demographic information, presenting complaint, associated ocular and systemic symptoms, past treatment history for ocular or systemic illnesses were taken. Socio-demographic data and relevant ophthalmic history was filled into the pretested structured questionnaire.

Presenting visual acuity; intraocular pressure measurement, slit-lamp examination of the anterior and posterior segment was performed. Posterior segment evaluation was performed with +90D funduscopic lens on the slit-lamp (slit-lamp biomicroscopy with 90D lens) for each patient by a vitreoretinal specialist after dilation of the pupil with 1% tropicamide eye drop. Indirect ophthalmoscopy with scleral indentation and +20D lens was used to examine the retinal periphery of all patients. B-scan ultrasound was used to examine the retina in patients with inadequate retinal view.

Grading of presenting visual acuity for visual impairment and blindness was done according to the WHO classification system as follows: visual acuity better or equal to 6/18 – normal; visual acuity between 6/24 and 6/60 – moderate visual impairment; visual acuity less than to 6/60 and better than or equal to counting fingers at 3m – severe visual impairment; visual acuity less than counting fingers at 3m – blindness; for bilateral blindness the result of the eye with the better presenting visual acuity was recorded.

### Data Analysis

The collected data was checked for accuracy, coded and entered in to SPSS Version 20 and analysis done; P value of less than 0.05 was considered as statistically significant.

### Ethical Clearance

The study was conducted in full compliance with the 1964 Helsinki declaration on research involving human subjects. Prior to commencement of the study, ethical clearance was obtained from the Ethics Committee (Institutional Review Board) of University of Gondar. Written informed consent was obtained from each study participant.

## Results

In this study, 65 consecutive new patients (73eyes) with retinal detachments participated. The median age of the participants was 49 with inter-quartile range of 38.5 years. Majority of participants 64.6% (42/65) were males. (Table 1). Majority of the participants, 80% (52/65), presented with complaint of visual reduction followed by complaints of visual reduction and floaters 12.3% (8/65).

**Table1:**
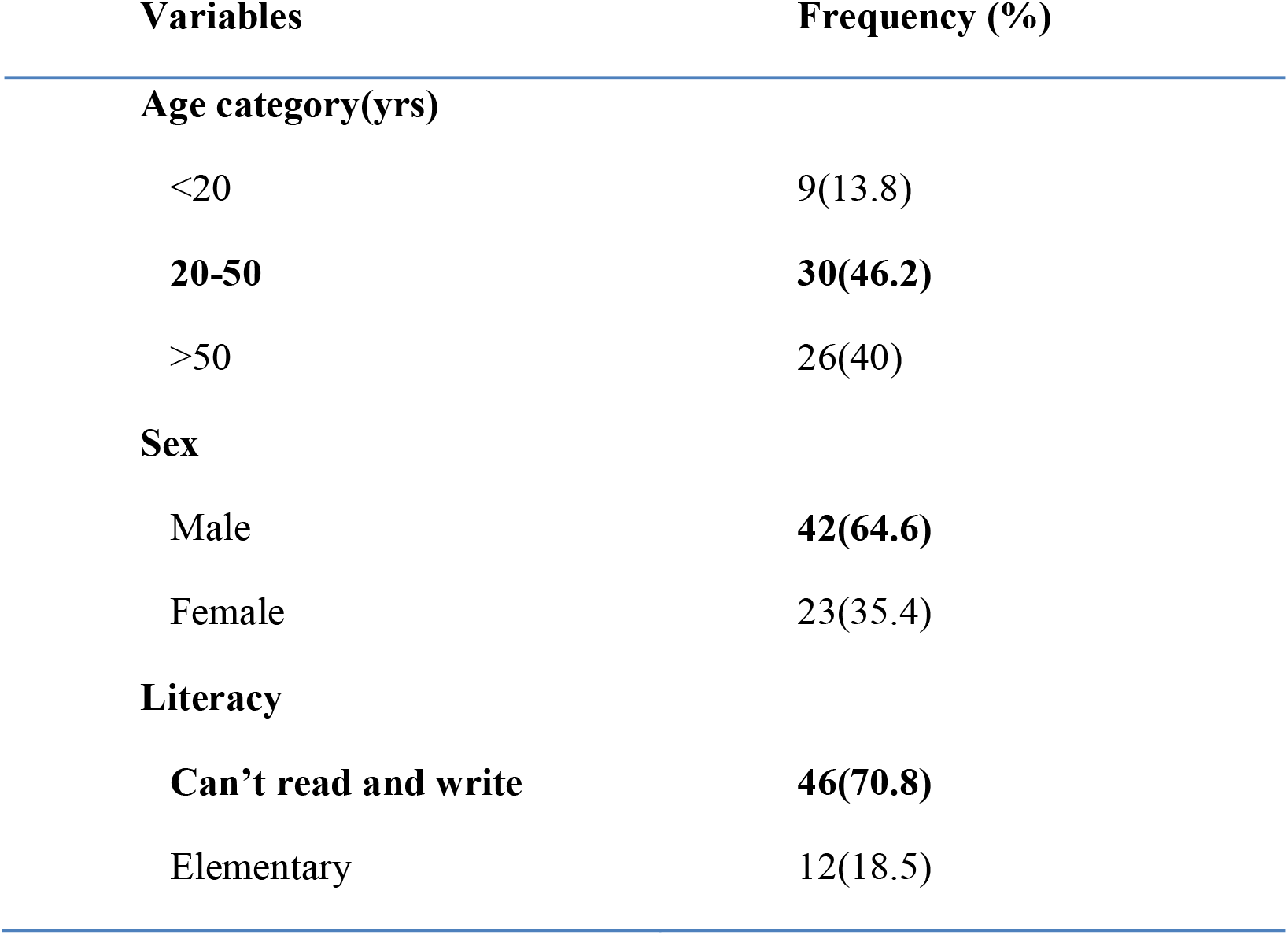
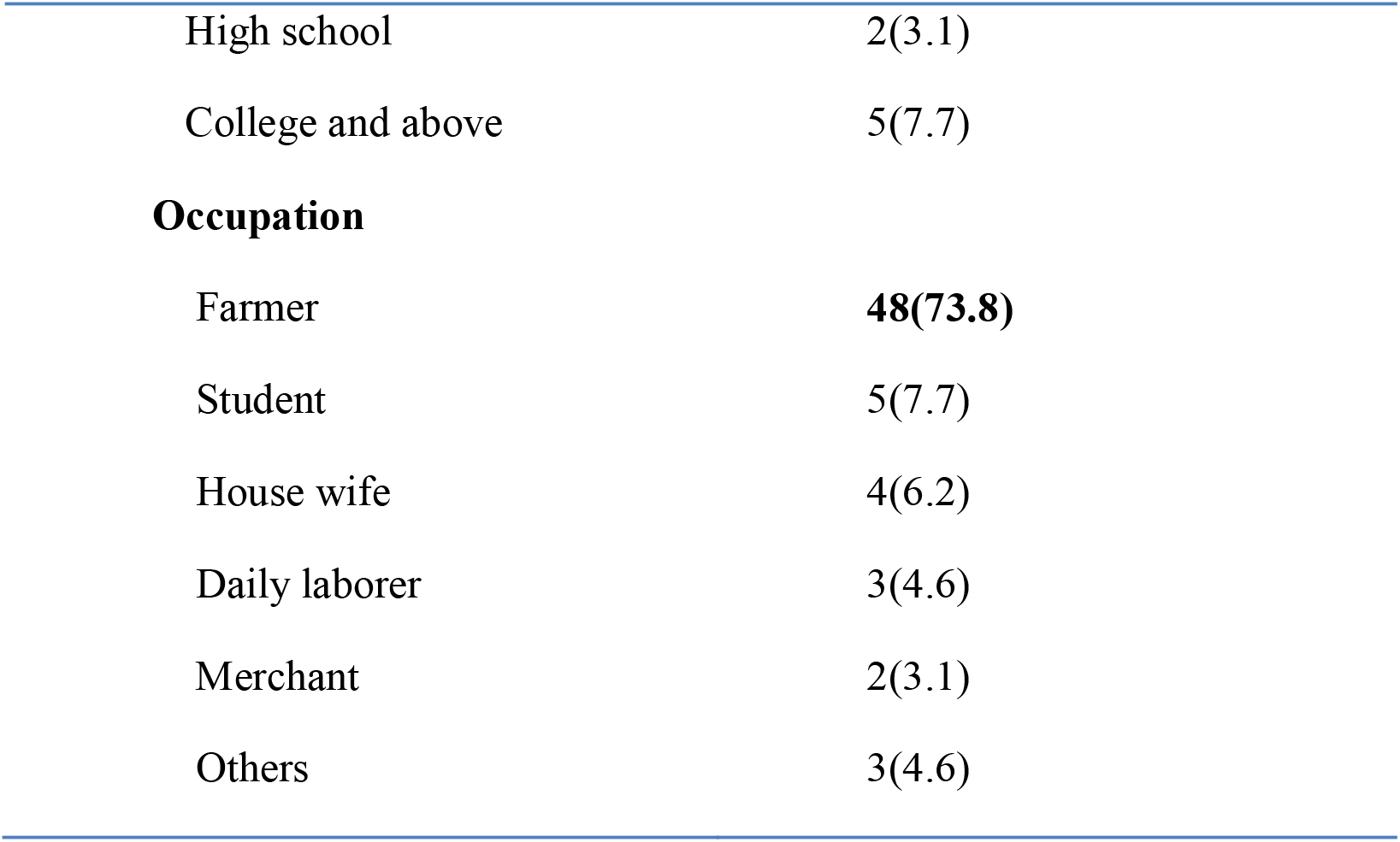
Socio-demographic characteristics of patients with retinal detachment at University of Gondar, North West Ethiopia, 2018 (n=65)

More than three quarter of the patients, 75.38 %(49/65), came after 1 month of the symptom occurrence; out of which 41% (20/49) visited the center after 1year duration.

Regarding the visual status of the participants; 92.3% (60/65) were blind at presentation in the affected eye and 27.7% (18/65) were bilaterally blind. Most of the participants had retinal detachment in only one eye, 87.69% (57/65). Ocular co-morbidities were found in 49.23% (32/65) of the participants; of which cataract being the most common one 53.13 %(17/32).(Table-2)

**Table2.**
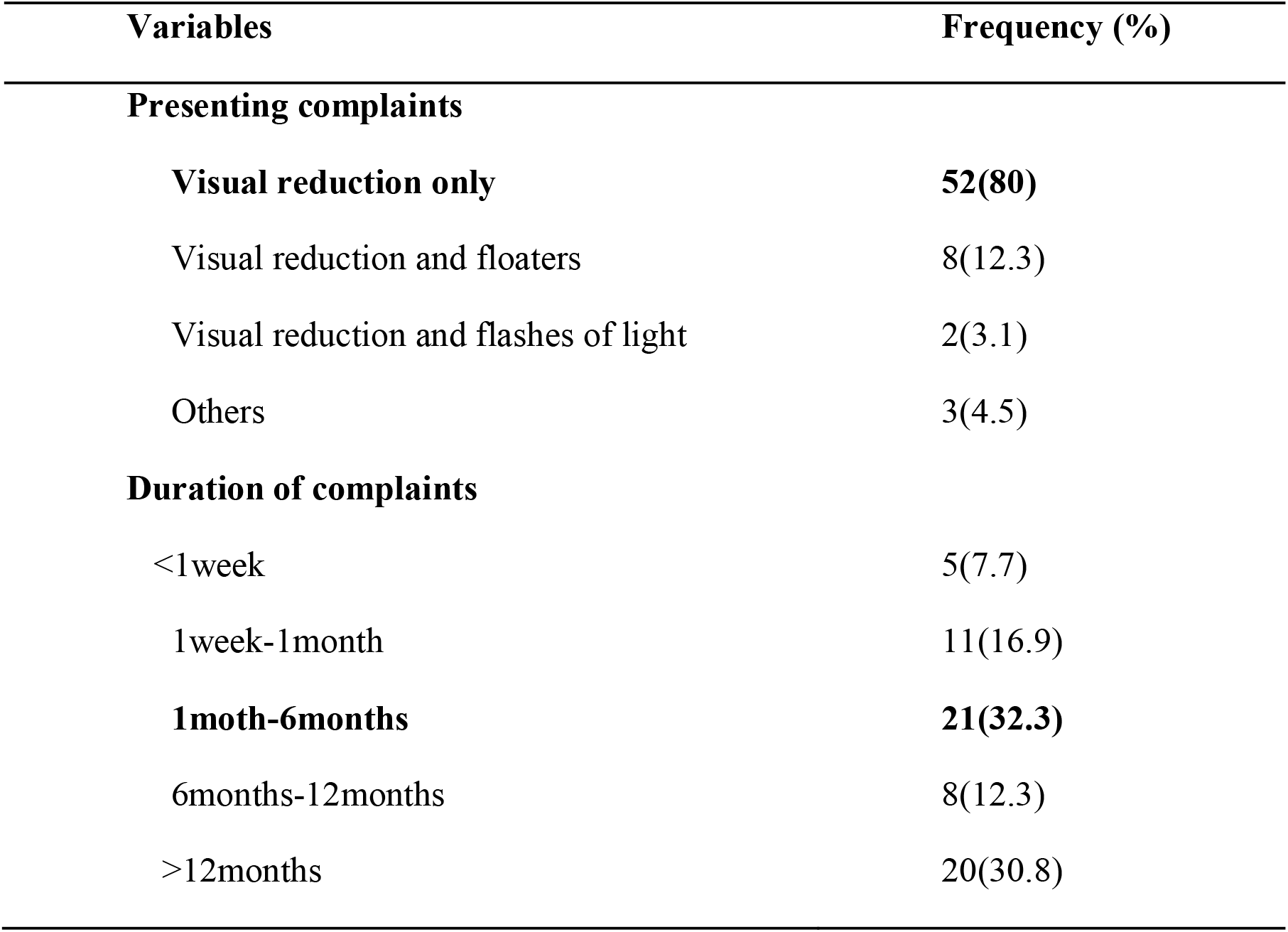
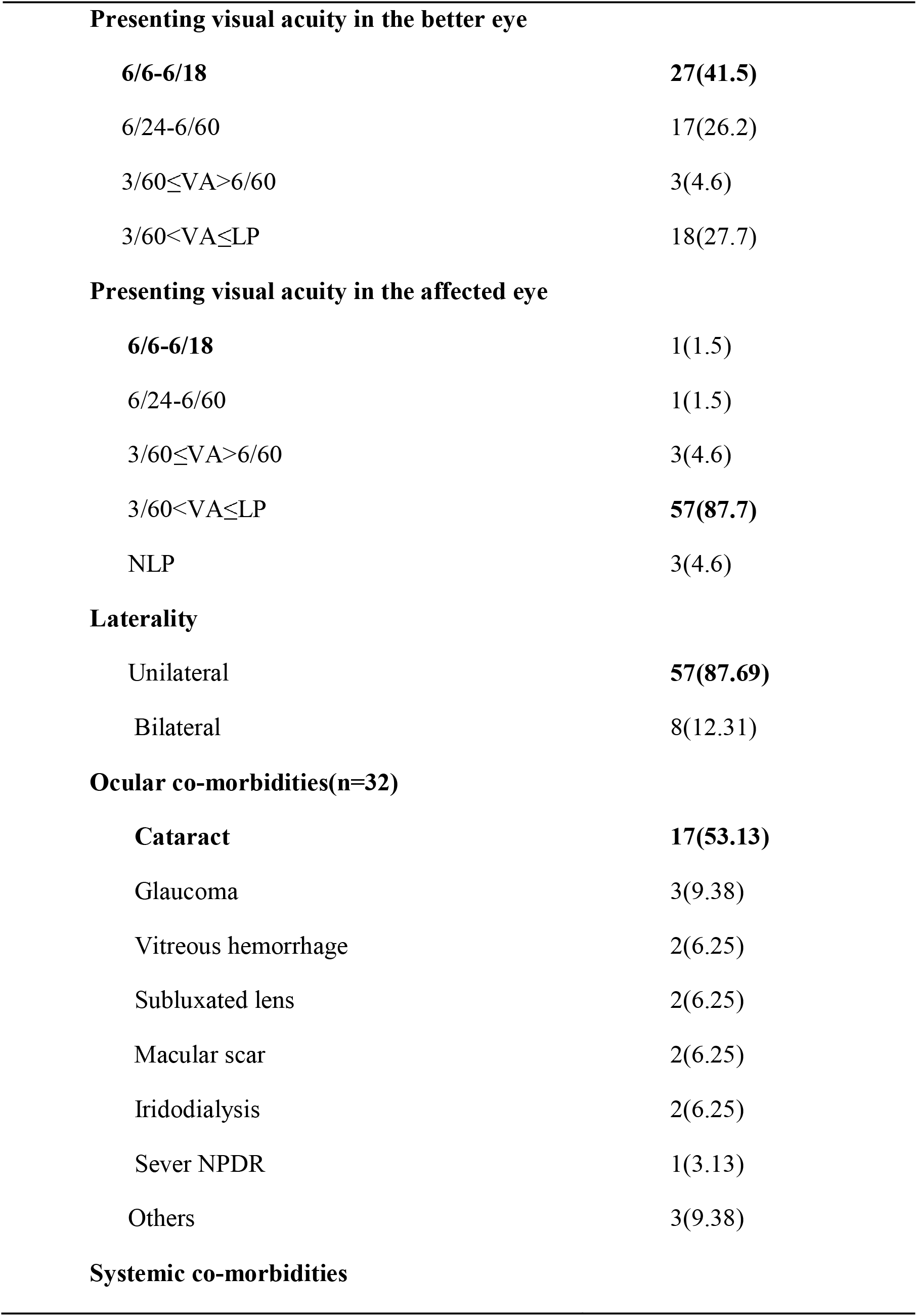
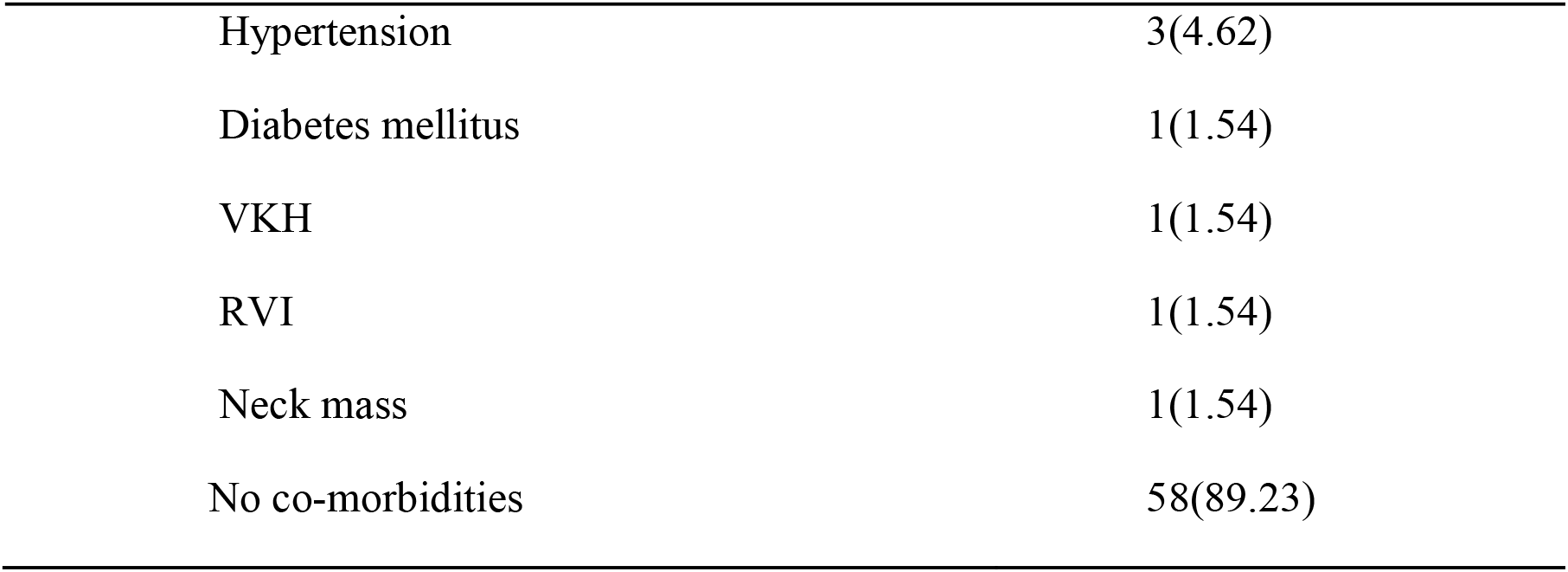
Clinical characteristics of patients with retinal detachment in University of Gondar, 2018 (n=65)

| **Variables** | **Frequency (%)** |
| --- | --- |
| **Presenting complaints** |  |
| **Visual reduction only** | **52(80)** |
| Visual reduction and floaters | 8(12.3) |
| Visual reduction and flashes of light | 2(3.1) |
| Others | 3(4.5) |
| **Duration of complaints** |  |
| <1week | 5(7.7) |
| 1 week-1month | 11(16.9) |
| **1moth-6months** | **21(32.3)** |
| 6months-12months | 8(12.3) |
| >12months | 20(30.8) |
| **Presenting visual acuity in the better eye** |  |
| **6/6-6/18** | **27(41.5)** |
| 6/24-6/60 | 17(26.2) |
| 3/60≤VA>6/60 | 3(4.6) |
| 3/60<VA≤LP | 18(27.7) |
| **Presenting visual acuity in the affected eye** |  |
| **6/6-6/18** | 1(1.5) |
| 6/24-6/60 | 1(1.5) |
| 3/60≤VA>6/60 | 3(4.6) |
| 3/60<VA≤LP | **57(87.7)** |
| NLP | 3(4.6) |
| **Laterality** |  |
| Unilateral | **57(87.69)** |
| Bilateral | 8(12.31) |
| **Ocular co-morbidities(n=32)** |  |
| **Cataract** | **17(53.13)** |
| Glaucoma | 3(9.38) |
| Vitreous hemorrhage | 2(6.25) |
| Subluxated lens | 2(6.25) |
| Macular scar | 2(6.25) |
| Iridodialysis | 2(6.25) |
| Sever NPDR | 1(3.13) |
| Others | 3(9.38) |
| **Systemic co-morbidities** |  |
| Hypertension | 3(4.62) |
| Diabetes mellitus | 1(1.54) |
| VKH | 1(1.54) |
| RVI | 1(1.54) |
| Neck mass | 1(1.54) |
| No co-morbidities | 58(89.23) |

The commonest type of retinal detachment was Rhegmatogenous Retinal Detachment (RRD), 89.23% (58/65), followed by Tractional Retinal Detachment (TRD); 6.15% (4/65). Retinal break was identified in 50 %(29/58) of the participants with RRD. Horse shoe shaped tear was the commonest break type; 48.28 %(14/29) and the supero-temporal retina was the commonest site of break, 10.34% (6/58). Most of the patients with RRD, 70.68% (41/58), had PVR of which 85.37 %(35/41) was grade C. Majority of the participants came with total retinal detachments, 55.4% (36/65), followed by inferior, 32.3 %(21/65) detachment. The macula was detached in 81.5% (53/65) of the participants at presentation (Table 3).

**Table 3:**
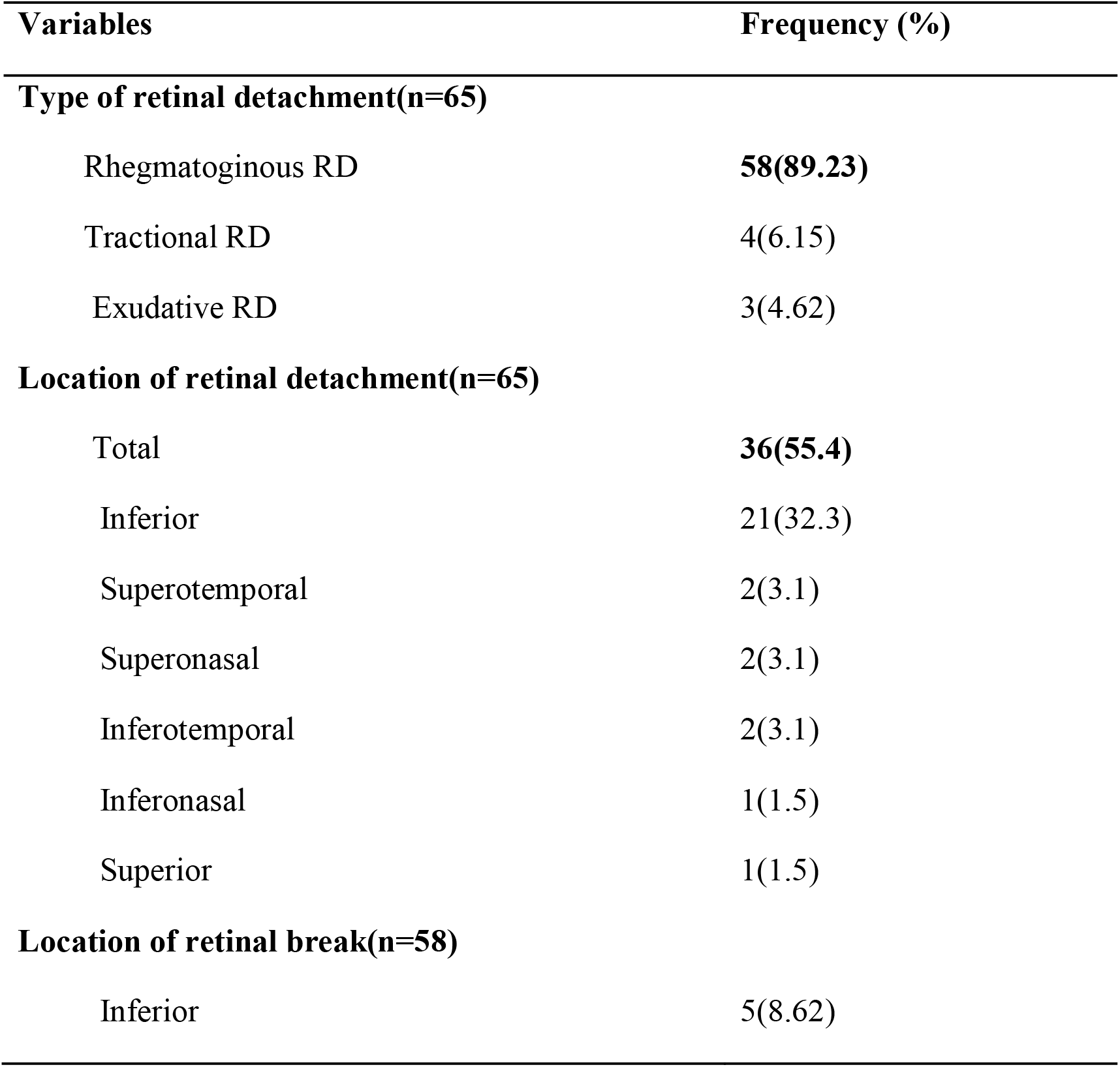
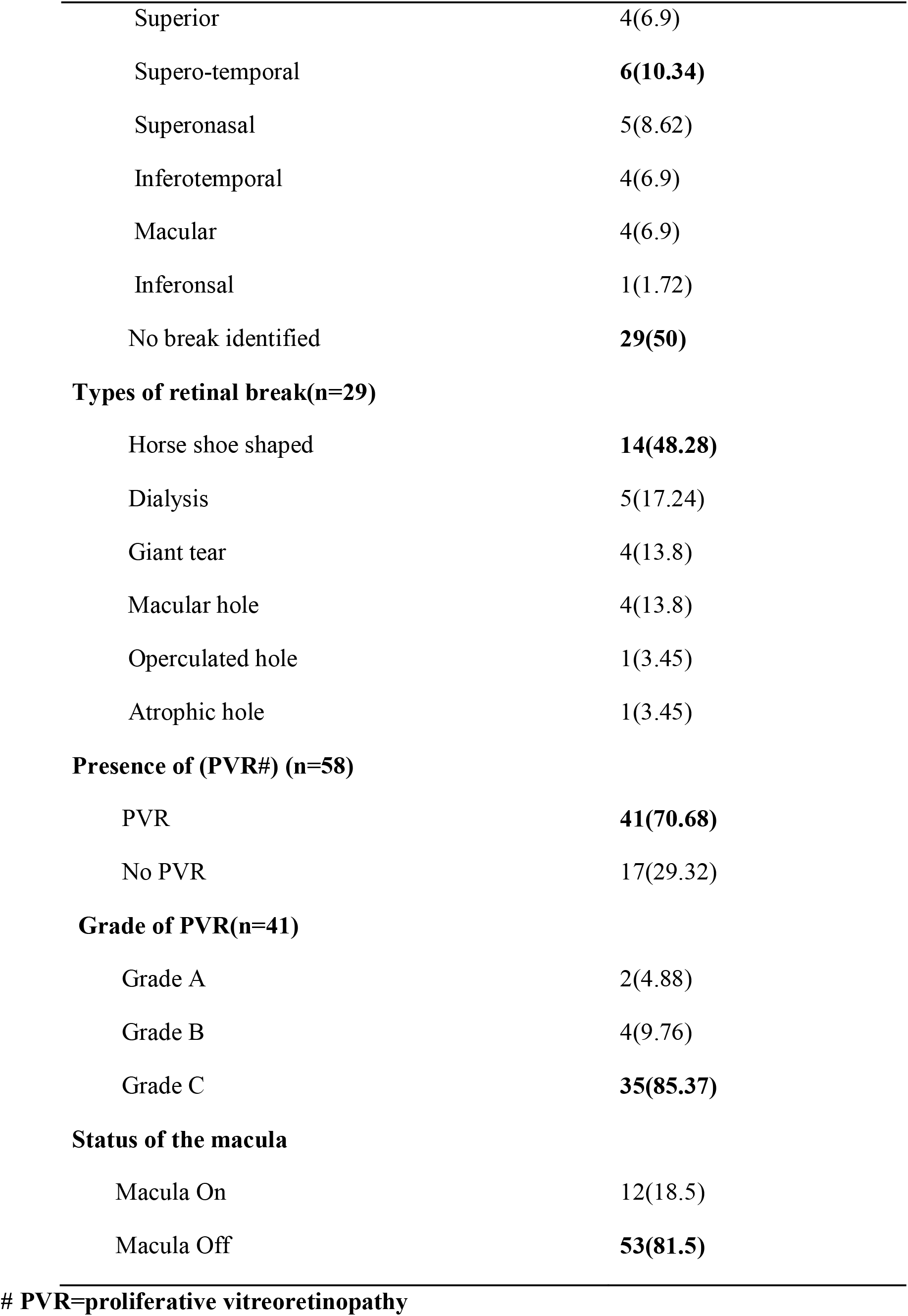
Patterns of retinal detachment among studied patients in University of Gondar; 2018 (n-65).

| **Variables** | **Frequency (%)** |
| --- | --- |
| **Type of retinal detachment(n=65)** |  |
| Rhegmatogenous RD | **58(89.23)** |
| Tractional RD | 4(6.15) |
| Exudative RD | 3(4.62) |
| **Location of retinal detachment(n=65)** |  |
| Total | **36(55.4)** |
| Inferior | 21(32.3) |
| Superotemporal | 2(3.1) |
| Superonasal | 2(3.1) |
| Inferotemporal | 2(3.1) |
| Inferonasal | 1(1.5) |
| Superior | 1(1.5) |
| **Location of retinal break(n=58)** |  |
| Inferior | 5(8.62) |
| Superior | 4(6.9) |
| Supero-temporal | **6(10.34)** |
| Superonasal | 5(8.62) |
| Inferotemporal | 4(6.9) |
| Macular | 4(6.9) |
| Inferonsal | 1(1.72) |
| No break identified | **29(50)** |
| **Types of retinal break(n=29)** |  |
| Horse shoe shaped | **14(48.28)** |
| Dialysis | 5(17.24) |
| Giant tear | 4(13.8) |
| Macular hole | 4(13.8) |
| Operculated hole | 1(3.45) |
| Atrophic hole | 1(3.45) |
| **Presence of (PVR#) (n=58)** |  |
| PVR | **41(70.68)** |
| No PVR | 17(29.32) |
| **Grade of PVR(n=41)** |  |
| Grade A | 2(4.88) |
| Grade B | 4(9.76) |
| Grade C | **35(85.37)** |
| **Status of the macula** |  |
| Macula On | 12(18.5) |
| Macula Off | **53(81.5)** |

Majority of the cases with delayed presentation had macular detachment (84%). Significant association between the presence of Proliferative Vitreoretinopathy (PVR) and the macular detachment was noted (p<0.001) where 95% of patients with PVR had detached macula (Table 4).

**Table 4:** Risk factors identified among participants with RD in University of Gondar; North West Ethiopia; 2018 (n=65)

| **Variables** | **Frequency (%)** |
| --- | --- |
| **Trauma** | **19(29.2)** |
| Pseudophakia | **11(16.9)** |
| Pathologic myopia | 6(9.2) |
| Posterior Uveitis | 6(9.2) |
| Lattice | 2(3.1) |
| PVD | 2(3.1) |
| Aphakia | 1(1.5) |
| No identified risk(idiopathic) | 18(27.7) |

Risk factors were identified in 72.31% (47/65) of the patients and trauma accounts for 40.43 % (19/47) of the identified risk factors followed by pseudophakia, 23.4% (11/47) and pathologic myopia, 9.2 %.(Table 4).

## Discussion

Rhegmatogenous retinal detachment was the commonest type of RD found in 89.23% patients (95% CI; 81% to 96%). About 71% of RRD cases had PVR. The most frequently noted break was horse-shoe tear (48%). The main identified risk factors was trauma (29%) followed by pseudophakia (17%). More than three-quarter of the patients (82%) had detached macula. Among 73 eyes presented with RD, 92.3% of them were blind.

The magnitude of RRD found in the present study is comparable with a study done in Nigeria, in which the prevalence was 93.6%. However, this is lower than what was reported at Jimma, South West Ethiopia (58.5%). The discrepancy might be due to our small sample size. (2,9)

Among patients with rhegmatogenous retinal detachment, 71% of them had PVR which is comparable with the reports in Jimma, Ethiopia (where 69 % had grade C PVR); but more than twice higher than the result in India (33%) This is likely to be due to the delayed presentation of our patients; in which more than 75% patients came after 1month as compared to less than 50% in the later one. (2,10).

Trauma (29.2%) was the commonest risk factor identified in this study. This is comparable with other previous reports from developing countries like South Africa (29.5%), Nigeria (35.5%) and Jimma, Ethiopia (34%) (11, 5, 2). This finding is also supported by the occupation of the participants, in which about three-quarter of the cases was farmers.

The second common risk factor identified was Pseudophakia (16.9%), which is comparable with the report at Menilik II, Ethiopia (14.2%) (9). However, this is lower than other reports from, Iran (32.2%), India (33.3-40%) and Pakistan (23.4%) (6, 12, 10, 13). This difference probably comes from the difference in the annual volume of cataract extraction.

High myopia was the third common risk factor (6.9%), identified in our participants. Though this result is comparable with the report in Nigeria (7.1%) (18/5), it’s much lower than in Jimma and Menilik II, Ethiopia (24.5% and 28.3% respectively), India (23.3%) and Iran (24%) (2, 9, 12, 6). This is likely to be from the fact that only participants with unexplained visual reduction in the fellow eye or those who had evidence of high myopia during examination were refracted.

For RRD cases, break was identified in 50% of participants and horse shoe tear was the commonest break (48.28%). Although this finding is similar with respect to type of tear, the proportion of identified break is lower than reports from Pakistan (68.2%), India (90%) and Jimma, Ethiopia (70%) (13, 10, 2).

The macula was detached in almost 82% of the patients in this study (95% CI; 74% to 90%). This is in agreement with the report at Jimma, Ethiopia (89.9%), but a little lower than Nigeria (91.5%) and Pakistan (93.5%) (2, 5, 13). However it is much higher than the reports in Olmsted County, Minnesota (41%) and Netherlands (54.5%) (14, 15). This could be due to the delayed presentation of patients in developing countries.

Majority of the eyes with retinal detachment (92.3%) were blind at presentation and 87.69% of the patients had unilateral retinal detachment. This is slightly higher than the results in Jimma, Nigeria, and India (64.5%, 79.8% and 53.3% respectively) (2,5, 12). This severe visual impairment in most of the patients also goes with the delayed presentation.

## Conclusions

The finding reveals that more than three-quarter of patients with retinal detachment visited our center after one month of symptom onset and majority of affected eyes were blind at presentation. Most of the participants had advanced PVR and detached macula. Trauma and pseudophakia were the two main risk factor identified. About 92% of eyes with RD were blind at presentation. To prevent the vision loss, facilities for surgical management of RD should be established at the center.

People should be educated to seek health care immediately after the onset of symptoms. Ocular safety precautions should be advised to decrease the risk of ocular trauma. Further study with large sample size is recommended

## Data Availability

Yes - all data are fully available without restriction

